# Individual-ecological triangulation of risk factors for peripheral artery disease: concordant evidence for smoking and diabetes, discordant evidence for kidney disease

**DOI:** 10.64898/2026.10.19.26365959

**Authors:** Ying Zhang, Jianjun Shi, Rong Wu, Fei Zhao

## Abstract

**Background:** Peripheral artery disease (PAD) is studied either through individual-level cohorts or national mortality statistics; the two evidence streams carry different biases and have not been formally compared within one design.

**Methods:** We triangulated two pre-specified streams. Individually, we analyzed 3,268 NHANES 1999-2004 participants aged 40 years and older with ankle-brachial index measurements (235 with PAD) linked to mortality through 2019, using survey-weighted logistic and Cox models, a Fine-Gray model, and attributable fractions. Ecologically, we used national Multiple Cause of Death records for 1999-2024 (390,174 PAD deaths) to estimate five-year-band trends in the share of PAD deaths mentioning tobacco (F17.0-F17.9), diabetes (E00-E89), or renal failure (N17-N19). Per a pre-specified protocol, we compared directions of association, each bridge quantified by a bootstrap direction-agreement probability.

**Results:** Weighted PAD prevalence was 5.05% (95% CI, 4.18 to 5.92). PAD was associated with cardiovascular death (hazard ratio, 2.65; 95% CI, 1.87 to 3.76). Smoking showed the strongest PAD association (odds ratio, 3.37) and the steepest rise in its mortality share (2.20% to 18.50%; direction-agreement probability, 1.000). Diabetes was associated with PAD (1.68) and rose after a 2014 trough (7.35% to 10.92%; probability, 1.000). Renal failure was associated with PAD (1.78) but its mortality share was flat for 26 years (14.55% to 14.49%; probability, 0.445).

**Conclusions:** The two streams agreed in direction for smoking and diabetes and diverged for kidney disease, reflecting different estimands. Smoking and diabetes are the factors supported by both streams for population-level PAD prevention.

## Introduction

Peripheral artery disease (PAD), atherosclerotic narrowing of the lower-extremity arteries, affects a substantial share of middle-aged and older adults and is strongly associated with loss of ambulation, amputation, and death [1]. Because PAD shares its risk factors, and much of its prognosis, with coronary and cerebrovascular atherosclerosis, it is increasingly treated as a marker of systemic atherosclerotic burden rather than an isolated limb condition [2], a framing embedded in contemporary national cardiovascular statistics [3]. Yet PAD remains underdiagnosed: population screening with the ankle-brachial index (ABI) repeatedly identifies large numbers of asymptomatic individuals who never reached clinical care [4]. How the leading risk factors translate into PAD at the level of individual patients, and into PAD deaths at the level of the whole population, are questions that the existing literature answers only separately.

The two dominant evidence streams rarely speak to each other. Individual-level studies link risk factors to prevalent or incident PAD in cohorts of finite size [5], with rich covariates but limited statistical power for mortality endpoints [6], and their findings depend on how comorbidities were measured at a single point in time [5]. Ecological studies, in contrast, describe large national series of deaths in which comorbidities appear as coded mentions, offering decades of coverage and essentially complete ascertainment of mortality, but no covariates [7], no individual exposure measurement, and well-known vulnerabilities to coding practice [8]. Reviews of PAD epidemiology cite both types of evidence interchangeably, although the two carry different biases, answer different questions, and have almost never been formally juxtaposed within a single design [9].

Triangulation offers a disciplined way to put them together [10]. Following that framework and recent planning guidance for triangulation studies [11], we specified, before examining the ecological data, the prior hypothesis that any risk factor positively associated with PAD at the individual level would also show a rising share of mentions among PAD deaths over time, on the reasoning that a growing risk-factor burden should reshape the comorbidity composition of national PAD mortality. We also pre-specified a conflict plan with three candidate explanations for a discordant result: differential measurement bias between the layers, compositional change unrelated to the risk factor, and cross-level confounding. Disagreement, when it appears, is therefore interpretable by design rather than treated as failure [10].

This study therefore proceeds in three steps. First, using the NHANES 1999-2004 ABI subsample linked to mortality through 2019, we quantify the association of five major risk factors, current smoking, diabetes, hypertension, hypercholesterolemia, and chronic kidney disease, with prevalent PAD, and the association of PAD itself with all-cause and cardiovascular death, with population attributable fractions for the leading factors.

Second, using 26 years of national Multiple Cause of Death records (1999-2024) with PAD as the underlying cause, we estimate time trends in the share of PAD deaths that also mention each of the three risk factors carried forward from the individual-level analysis, analyzed as three pre-specified comorbidity bridges. Third, we compare the direction of the individual-level and ecological findings within a pre-specified triangulation matrix, and interpret concordance and discordance under the pre-specified conflict plan.

## Methods

### Data sources and study design

This study used two complementary, non-overlapping evidence streams. The individual-level stream drew on the National Health and Nutrition Examination Survey (NHANES) 1999-2004 ankle-brachial index (ABI) fasting subsample, linked through 2019 to mortality records under the NCHS linkage program [12]. The ecological stream drew on the CDC WONDER Multiple Cause of Death databases: the 1999-2020 Bridged-Race file (D77) and the 2018-2024 Single-Race file (D157). The two streams were analyzed separately and then compared at the level of direction of association, following a pre-specified triangulation protocol [10]. Because both sources are publicly released, aggregated or de-identified datasets, the analysis did not require institutional review board approval; use of the linked mortality files followed the NCHS data use restrictions, and no cell based on fewer than 10 deaths is reported.

### Individual-level analysis population

Of 3,268 participants aged 40 years or older with valid ABI measurements in the 1999-2004 fasting subsample, 235 met the definition of peripheral artery disease (PAD), operationalized as ABI of 0.9 or lower in either leg. The analytic cohort contributed 1,245 deaths through December 31, 2019. Mortality follow-up used the public-use linked mortality files, which provide date and cause of death grouped into 10 leading-cause categories; disease-specific (ICD-coded) causes of death are not released at the individual level. Cardiovascular death was therefore defined as death from diseases of heart (UCOD_LEADING 001) or cerebrovascular disease (005). Follow-up time was computed from months of examination follow-up, censored administratively at the end of the linkage window.

### Measures

PAD was the primary outcome. Risk factors were defined as follows: diabetes as self-reported physician diagnosis or fasting plasma glucose of 126 mg/dL or higher; current smoking as self-reported ever smoking at least 100 cigarettes with current everyday or some-day smoking; hypertension as self-reported diagnosis or measured systolic blood pressure of 140 mmHg or higher; hypercholesterolemia as total cholesterol of 240 mg/dL or higher; and chronic kidney disease (CKD) as estimated glomerular filtration rate below 60 mL/min/1.73 m2 by the CKD-EPI 2009 equation [13], with serum creatinine unit-checked before computation. All survey analyses incorporated the 6-year fasting subsample weights with Taylor-series linearization, cycle-specific primary sampling units and strata, and a lonely-PSU adjustment option.

### Statistical analysis, individual level

We first estimated the weighted prevalence of PAD overall and by risk-factor stratum. We then fitted survey-weighted logistic models for PAD with each risk factor, first adjusted for age (per 10 years), sex, and race/ethnicity, then mutually adjusted for the other four risk factors. Survival analyses used survey-weighted cause-specific Cox proportional hazards models with PAD as the exposure and all-cause and cardiovascular death as outcomes, adjusted for age, sex, and race/ethnicity (primary model); additional adjustment for body mass index served as a sensitivity analysis and changed the PAD hazard ratio by 0.2% or less (all-cause 2.121 to 2.121; cardiovascular 2.652 to 2.646). Competing risks were addressed with a Fine-Gray subdistribution hazard model implemented through risk-set augmentation, because the standard competing-risks package does not accept survey weights. The population attributable fraction (PAF) of cardiovascular death due to PAD was computed with the Levin formula. Because PAF estimates are formula-dependent [14], we repeated all computations with the Miettinen formula; the two approaches agreed on the qualitative conclusion that smoking contributes one of the largest attributable shares but differed in magnitude by up to 12 percentage points, so no precise attribution ranking is claimed and both sets of estimates are provided in the supplementary material.

### Ecological analysis, comorbidity bridges

The denominator for each bridge was the annual count of deaths with PAD as the underlying cause of death, defined as ICD-10 I70.2 (atherosclerosis of arteries of the extremities) or I70.9 (generalized and unspecified atherosclerosis). Numerators were any-mention counts of the index comorbidity on the same death records. Three bridges were pre-specified on the basis of the individual-level findings: diabetes (chapter-level E00-E89), tobacco use (F17.0-F17.9), and renal failure (N17-N19 as the primary definition, matching the individual-level CKD construct; the broader N00-N99 chapter served as a sensitivity definition and produced the same conclusion). For each bridge we computed the annual share of PAD deaths mentioning the comorbidity, aggregated shares into 5-year bands, tested the first-versus-last band contrast with a two-proportion z test, and fitted a linear trend to the annual log-odds of the share. The 2018-2020 overlap between the two WONDER files was verified year by year; counts agreed exactly for every bridge, and the files were joined by taking D77 for 1999 through 2020 and D157 for 2021 through 2024. ICD-10 coding-rule changes effective in 2003 produced a discontinuity in F17 mentions (share rising from 1.0% to 3.3%), so the tobacco bridge is interpreted with caution and with emphasis on the post-2005 window.

### Triangulation protocol

Before analysis, we specified the prior hypothesis that a risk factor showing a positive individual-level association with PAD would also show a rising share of mentions among PAD deaths over time, on the reasoning that a growing risk-factor burden should reshape the comorbidity composition of PAD deaths. We also pre-specified a conflict plan with three candidate explanations for discordant findings: differential measurement bias between layers, compositional change unrelated to risk, and cross-level confounding. The comparison was restricted to direction of association; no cross-level causal inference is attempted. To make the directional comparison quantitative, each bridge was assigned a direction-agreement probability: the bootstrap probability that the share of mentions among PAD deaths was higher in the last (2020-2024) than in the first (2000-2004) five-year band, estimated by resampling calendar years within each band with replacement and pooling numerators and denominators over the sampled years (10,000 replicates).

Probabilities of at least 0.975 were classified as rising, at most 0.025 as falling, and values in between as flat; concordance with a positive individual-level association required a rising classification. For the tobacco bridge the probability was additionally computed over the post-2005 window because of the 2003 coding-rule change, and for diabetes from its 2010-2014 trough.

## Results

### Individual level: prevalence and risk factors for PAD

The weighted prevalence of PAD was 5.05% (95% CI, 4.18 to 5.92) among US adults aged 40 years and older in 1999-2004 (Figure 1). Prevalence rose markedly across every risk-factor stratum except hypercholesterolemia: from 4.34% to 9.60% with diabetes, from 4.42% to 7.41% with current smoking, from 2.77% to 7.71% with hypertension, and from 3.99% to 16.45% with CKD, the largest relative difference of the five factors (Table 1).

**Figure 1.**
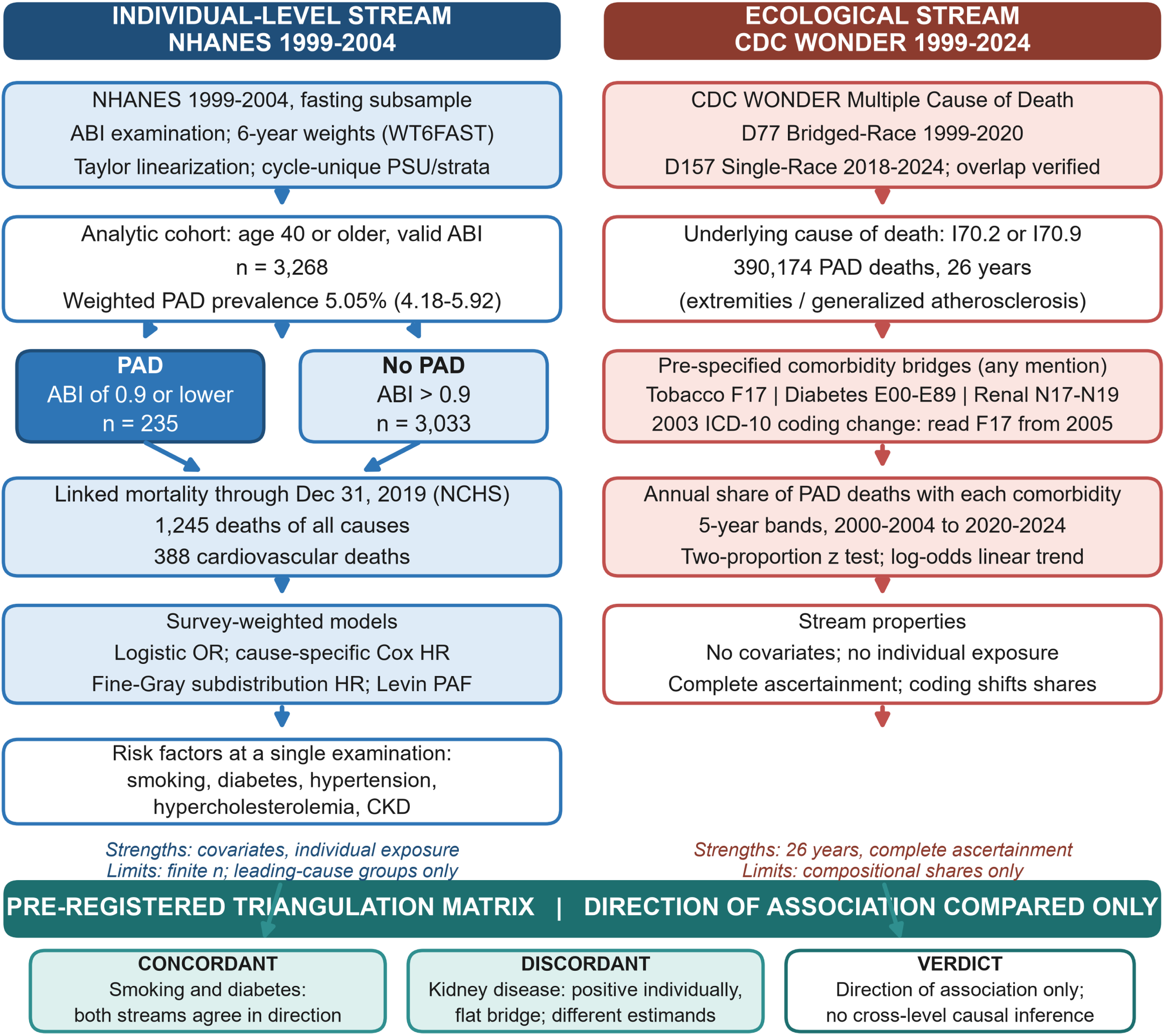
Study design and evidence flow. Two parallel, non-overlapping evidence streams were assembled: an individual-level stream (NHANES 1999-2004 ABI fasting subsample, 3,268 adults, 235 with PAD, linked to mortality through 2019) and an ecological stream (CDC WONDER Multiple Cause of Death, 390,174 deaths with PAD as the underlying cause, 1999-2024). Three pre-specified comorbidity bridges (tobacco, diabetes, renal failure) connect the streams in a pre-specified triangulation matrix with a pre-specified conflict plan; concordant, discordant, and overall verdict cards summarize the directional comparison.

**Table 1.**
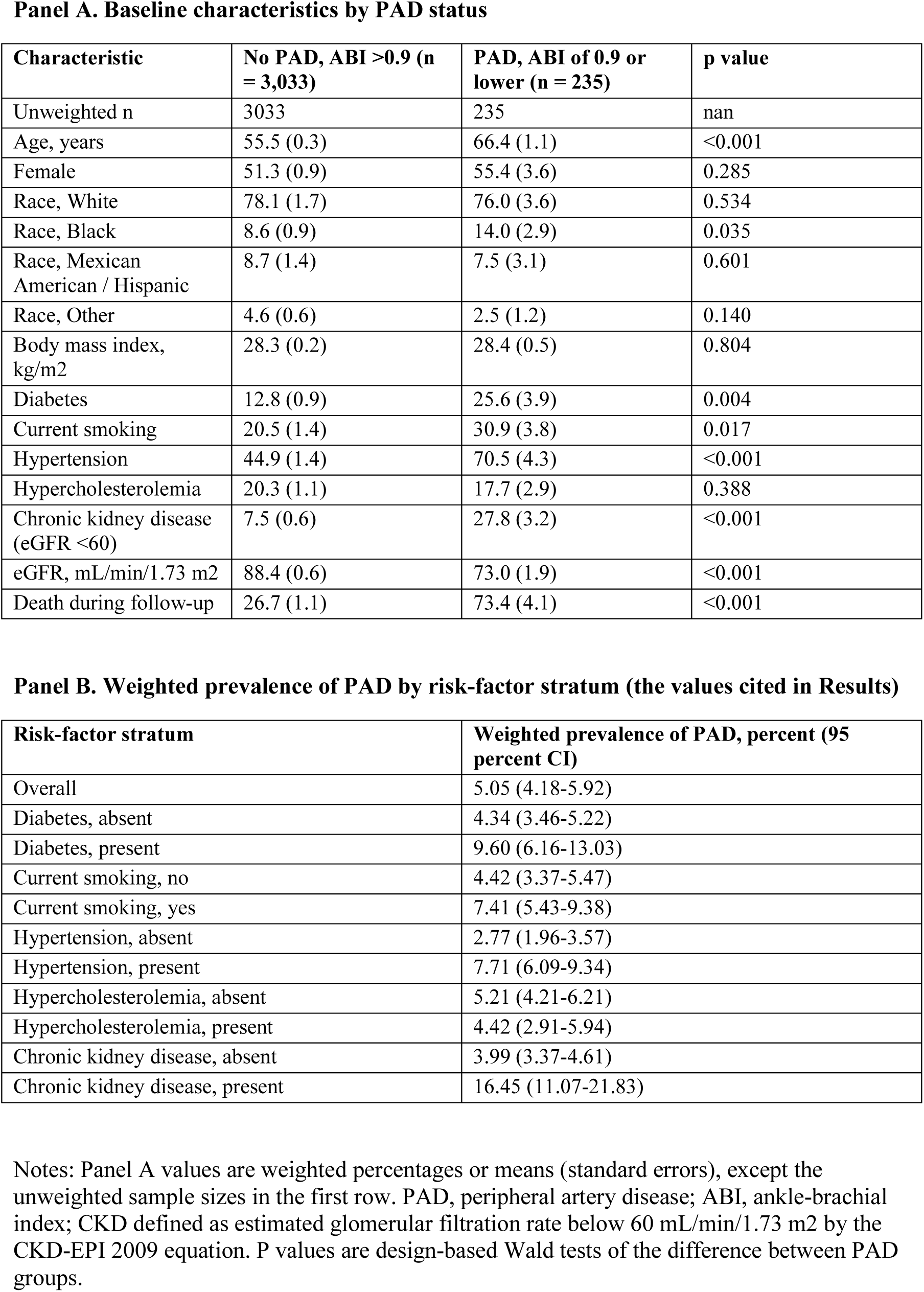
Weighted characteristics of the NHANES 1999-2004 ABI fasting subsample overall and by PAD status, with weighted PAD prevalence by risk-factor stratum.

In survey-weighted logistic models adjusted for age, sex, and race/ethnicity, current smoking (odds ratio [OR], 3.21; 95% CI, 1.98 to 5.23), CKD (OR, 1.83; 95% CI, 1.17 to 2.85), diabetes (OR, 1.90; 95% CI, 1.20 to 3.00), and hypertension (OR, 1.58; 95% CI, 1.02 to 2.43) were each associated with PAD, whereas hypercholesterolemia was not (OR, 0.83; 95% CI, 0.52 to 1.30). With mutual adjustment for all five factors, estimates were essentially preserved for the four positive factors: smoking 3.37 (2.05 to 5.56), CKD 1.78 (1.13 to 2.80), diabetes 1.68 (1.02 to 2.77), and hypertension 1.55 (0.99 to 2.42, overlapping unity) (Figure 3, Table 2).

**Table 2.**
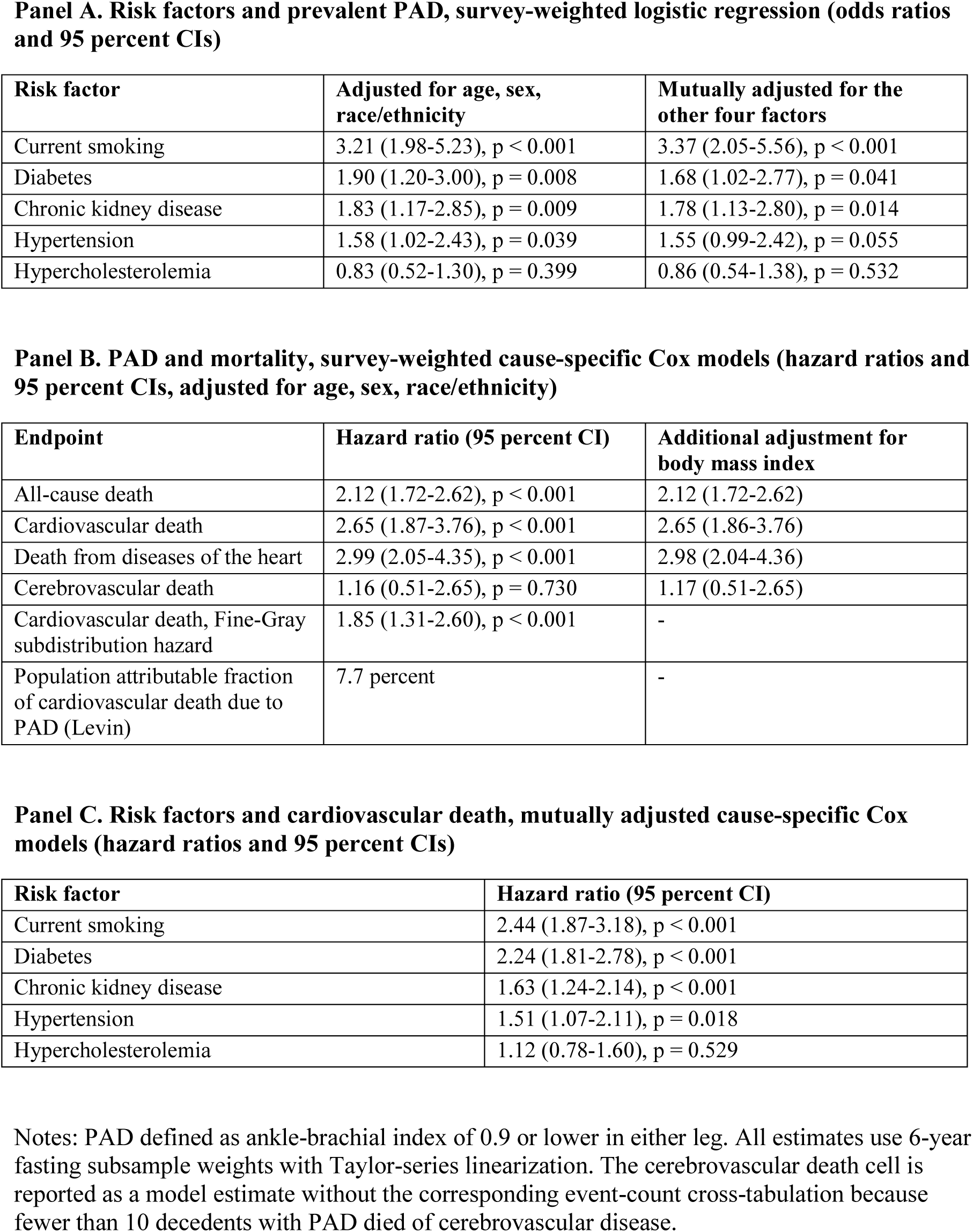
Survey-weighted models: (A) odds ratios for prevalent PAD; (B) hazard ratios for mortality associated with PAD, including Fine-Gray and sensitivity models; (C) hazard ratios for cardiovascular death by risk factor.

### Individual level: PAD and mortality

Over follow-up, 1,245 participants died, 388 of them from cardiovascular causes. PAD was associated with all-cause mortality (hazard ratio [HR], 2.12; 95% CI, 1.72 to 2.62) and, more strongly, with cardiovascular death (HR, 2.65; 95% CI, 1.87 to 3.76) and death from diseases of the heart (HR, 2.99; 95% CI, 2.05 to 4.35); the association with cerebrovascular death was absent (HR, 1.16; 95% CI, 0.51 to 2.65). Additional adjustment for body mass index changed none of these estimates by more than 0.2%.

Because noncardiovascular deaths compete with cardiovascular deaths, we refitted the cardiovascular endpoint as a Fine-Gray subdistribution hazard model; the subdistribution hazard ratio was 1.85 (95% CI, 1.31 to 2.60), attenuated but still well above unity. The Levin population attributable fraction of cardiovascular death due to PAD was 7.7%.

The same risk factors predicted cardiovascular death among these adults: in mutually adjusted cause-specific Cox models, smoking (HR, 2.44; 95% CI, 1.87 to 3.18), diabetes (HR, 2.24; 95% CI, 1.81 to 2.78), CKD (HR, 1.63; 95% CI, 1.24 to 2.14), and hypertension (HR, 1.51; 95% CI, 1.07 to 2.11) each raised cardiovascular mortality, while hypercholesterolemia again did not (HR, 1.12; 95% CI, 0.78 to 1.60).

Attributable fractions for PAD itself, computed with the Levin formula, identified current smoking as the dominant contributor (PAF, 33.3%), followed by hypertension (20.2%), diabetes (8.4%), and CKD (6.3%); the Miettinen formulation gave the same qualitative picture with different magnitudes and is reported in the supplementary material.

### Ecological level: comorbidity bridges among PAD deaths, 1999-2024

The national series comprised 390,174 deaths with PAD as the underlying cause across 1999-2024 (Figure 1). Each pre-specified bridge behaved differently (Figure 2, Table 3).

**Figure 2.**
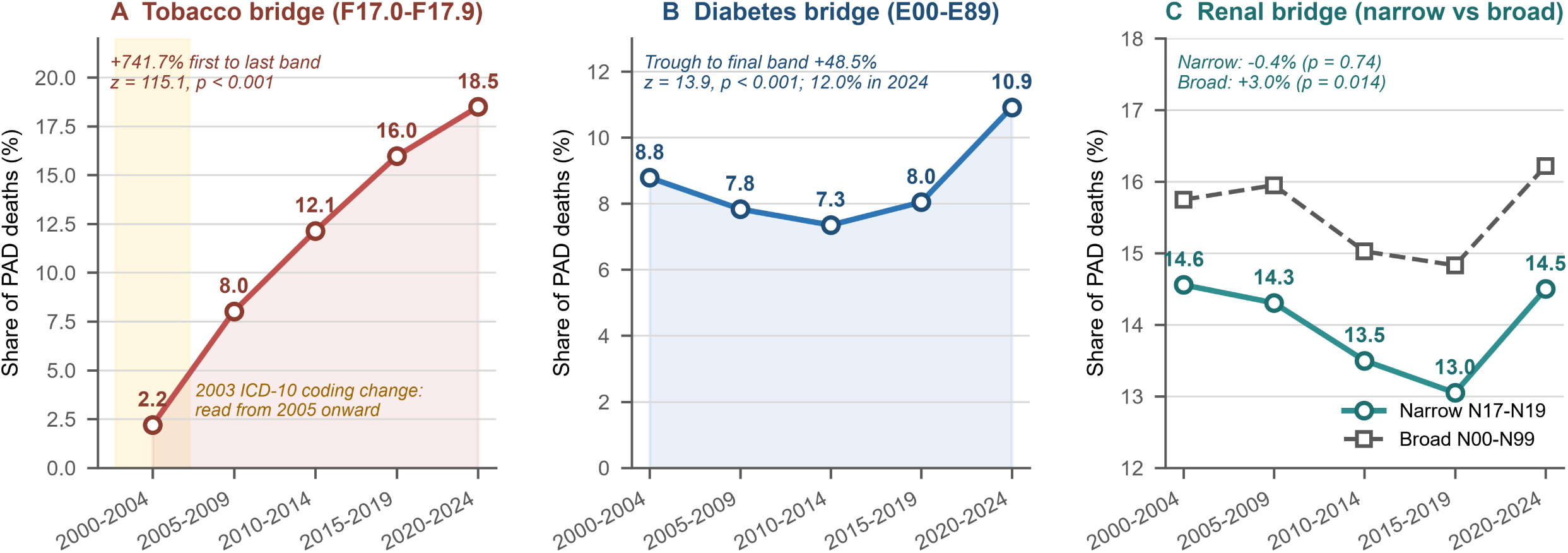
Comorbidity bridges among deaths with PAD as the underlying cause, United States, 1999-2024. Five-year-band shares of PAD deaths mentioning (A) tobacco use (F17.0-F17.9), (B) diabetes (E00-E89), and (C) renal failure (N17-N19, narrow definition) with the broader N00-N99 chapter as a sensitivity definition. The amber highlight marks the segment affected by the 2003 ICD-10 coding-rule change in the tobacco bridge.

**Figure 3.**
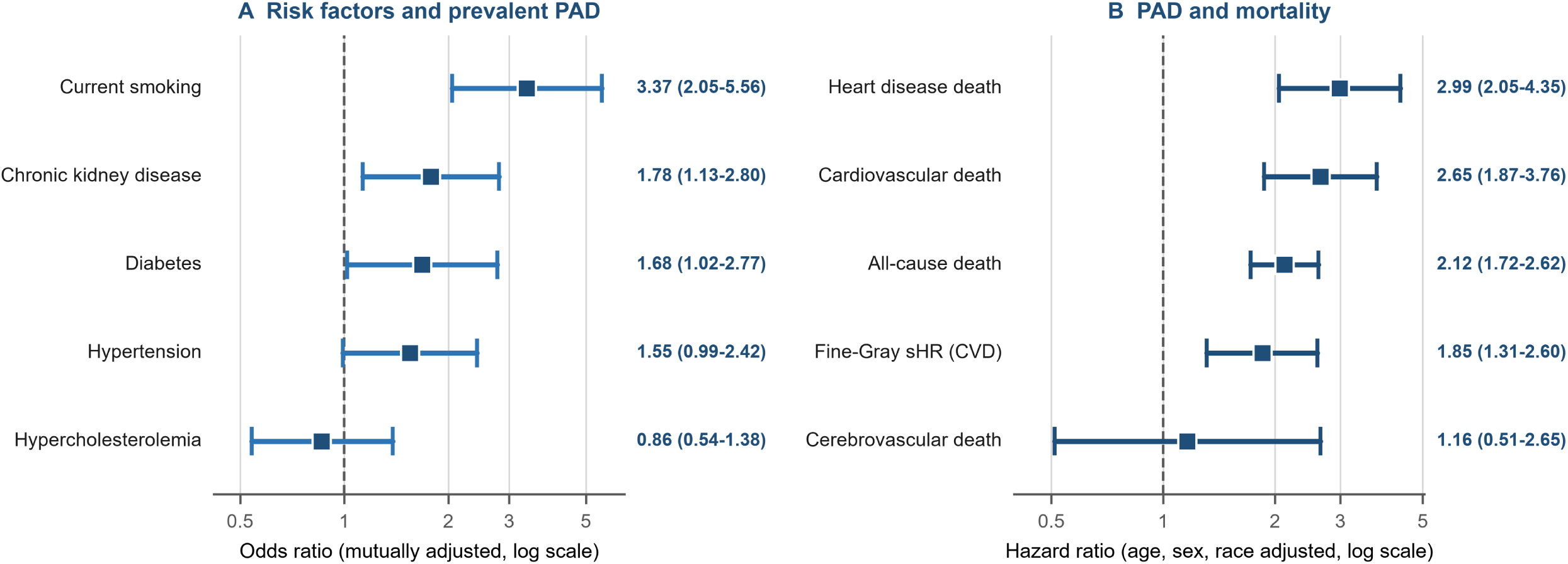
Individual-level associations from the NHANES 1999-2004 ABI subsample (survey-weighted, log scale). (A) Odds ratios for prevalent PAD, mutually adjusted for the other four risk factors. (B) Hazard ratios for mortality associated with PAD, adjusted for age, sex, and race/ethnicity, including the Fine-Gray subdistribution hazard ratio for cardiovascular death. Point estimates and 95% confidence intervals are labeled.

**Table 3.**
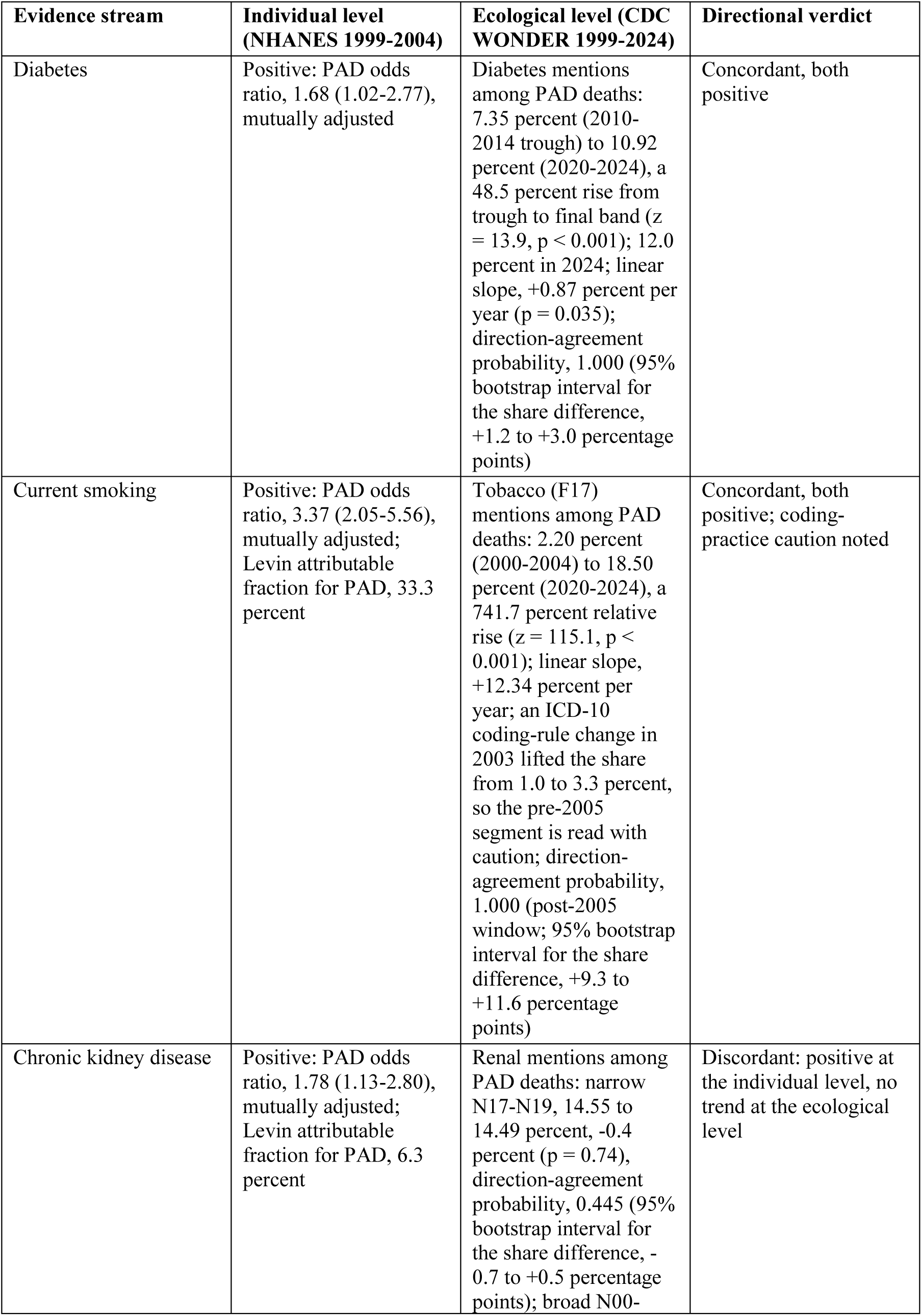
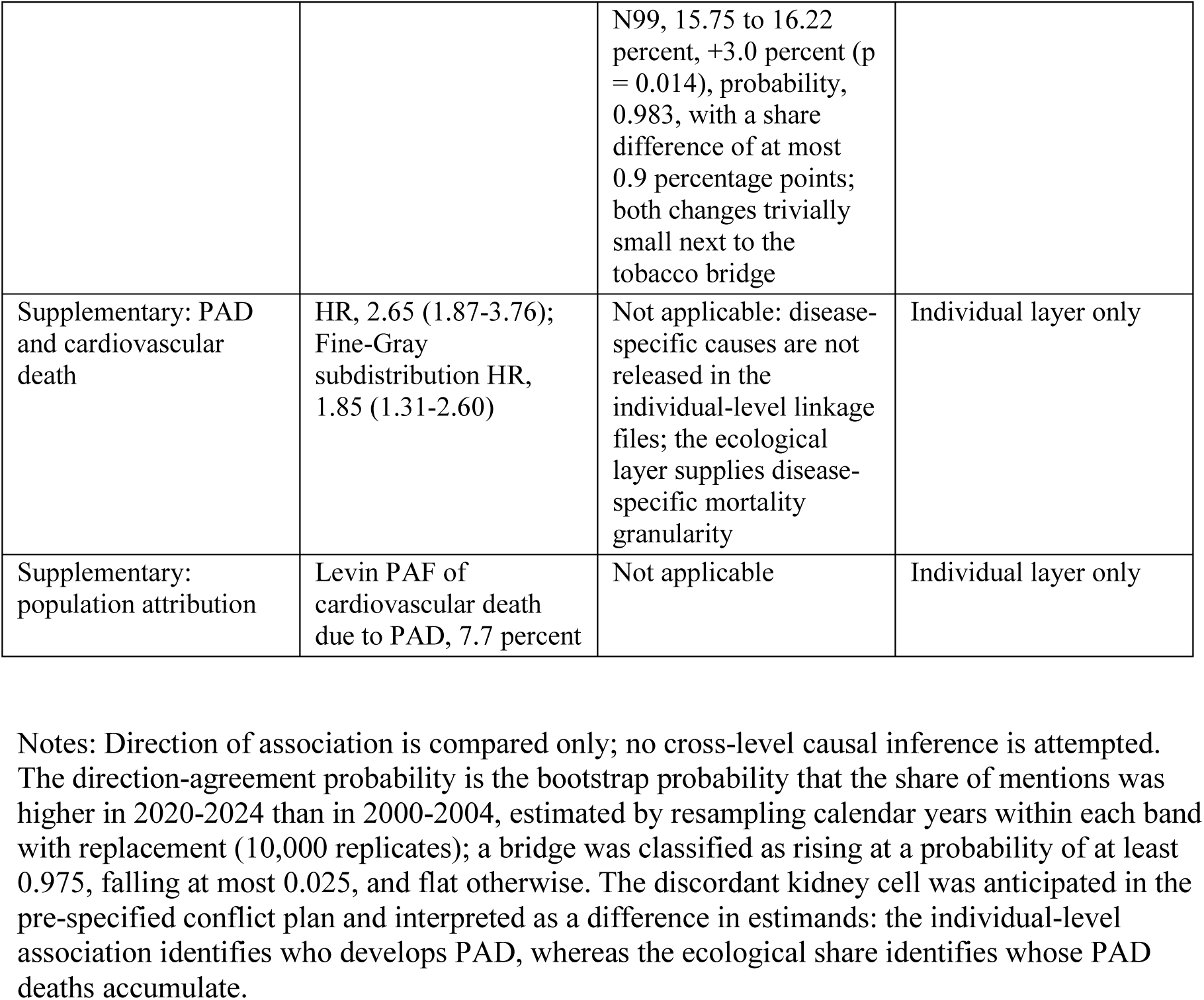
Pre-specified triangulation matrix: individual-level versus ecological directions of association for the three bridges.

The tobacco bridge (any mention of F17.0-F17.9) rose monotonically and steeply: from 2.20% of PAD deaths in 2000-2004 to 8.01% in 2005-2009, 12.15% in 2010-2014, 15.97% in 2015-2019, and 18.50% in 2020-2024, a relative increase of 741.7% (z = 115.1, p < 0.001); the annual log-odds slope was +12.34% per year (p < 0.001). Because an ICD-10 coding-rule change in 2003 lifted the share from 1.0% to 3.3%, the pre-2005 segment understates tobacco mentions, and the trend is most securely read from 2005 onward, over which it continued to climb without reversal.

The diabetes bridge (any mention of E00-E89) followed a U-shaped course: 8.78% in 2000-2004, falling to a trough of 7.35% in 2010-2014, then rising to 10.92% in 2020-2024, a relative increase of 48.5% from trough to final band (z = 13.9, p < 0.001), reaching 12.0% in 2024.

The renal-failure bridge (any mention of N17-N19) was flat across the entire 26-year window: 14.55% in 2000-2004 versus 14.49% in 2020-2024, a change of -0.4% (z = - 0.33, p = 0.74), with an annual log-odds slope of -0.17% per year (p = 0.40). Using the broader N00-N99 chapter instead gave a near-identical picture (15.75% to 16.22%, +3.0%, p = 0.014), an order of magnitude smaller than the tobacco-bridge rise and stable in every band, and the narrow and broad definitions tracked each other within 2 percentage points throughout, with renal failure accounting for 88% to 92% of chapter mentions throughout.

### Triangulation matrix

Directional comparison of the two streams (Table 3) yielded concordant findings for two of the three pre-specified bridges. Smoking showed the strongest individual-level association with PAD (OR, 3.37) and, in parallel, the steepest rise in its share of PAD deaths (direction-agreement probability, 1.000; 95% bootstrap interval for the share difference, +9.3 to +11.6 percentage points over the post-2005 window). Diabetes showed a positive individual-level association (OR, 1.68) and a rising ecological share after 2014 (probability, 1.000; 95% bootstrap interval, +1.2 to +3.0 percentage points). Kidney disease diverged: positively associated with PAD at the individual level (OR, 1.78), yet flat as a share of PAD deaths over 26 years under the primary narrow definition (probability, 0.445; 95% bootstrap interval, -0.7 to +0.5 percentage points), with the broad N00-N99 sensitivity definition directionally positive but quantitatively trivial (probability, 0.983; share difference of at most 0.9 percentage points). Under the pre-specified conflict plan, this discordance reflects a difference in estimands rather than a contradiction: the individual-level association identifies who develops PAD, whereas the ecological share identifies whose PAD deaths accumulate over time, and the flat renal-failure bridge argues against worsening kidney comorbidity as a driver of the national PAD mortality trend.

## Discussion

### Principal findings

In this study we brought together two evidence streams that are usually cited side by side but rarely examined together: individual-level data from a nationally representative examination survey, and 26 years of national mortality records. Three findings organize the results. First, at the individual level, PAD carried more than a twofold higher risk of all-cause death and a near-threefold risk of cardiovascular death, and among the five risk factors examined, current smoking, diabetes, chronic kidney disease, and hypertension were each independently associated with prevalent PAD, while hypercholesterolemia was not. Second, at the ecological level, the comorbidity composition of national PAD deaths moved in different directions for different factors: tobacco mentions rose almost without interruption across a quarter century, diabetes mentions followed a U-shaped course that has been climbing since 2014, and renal-failure mentions stayed flat within one to two percentage points for the entire window. Third, the two streams agreed in direction for smoking and diabetes and diverged for kidney disease, a pattern that was anticipated in the pre-specified conflict plan and is interpretable rather than merely inconvenient.

### The smoking and diabetes bridges: concordant by construction, informative by magnitude

For smoking, the concordance ran deeper than direction. The individual-level association was the strongest of the five factors (OR, 3.37), the Levin attributable fraction placed smoking among the largest contributors to PAD prevalence, and the national bridge rose by more than sevenfold in relative terms between the first and last five-year bands.

Cohort evidence likewise ties current smoking to major cardiovascular and limb events in established PAD [15], and newer lines of work, including smoking-sensitive mosaic loss of chromosome Y and its link to symptomatic disease, continue to reinforce the individual-level association [16]. The two layers, however, do not merely repeat each other: the cohort estimate carries the covariate control and the individual exposure measurement, while the national series shows the same factor visibly reshaping the composition of PAD deaths across generations of decedents. The concordance is therefore an example of what triangulation is designed to detect, evidence streams with different bias structures pointing the same way [10].

Two cautions attach to this reading. The 2003 ICD-10 coding-rule change lifted the tobacco share from 1.0% to 3.3%, so the early segment of the series understates mentions and the trend is most securely read from 2005 onward; over that later window the climb continued without reversal, which is why we retain the directional conclusion while flagging the artifact. The population harms of smoking across the life course remain among the largest of any modifiable exposure [17]. For diabetes, the U-shaped trajectory is itself a finding: a falling share through 2014 followed by a sustained rise to 10.92% in 2020-2024, with 2024 alone at 12.0%. The individual-level association (OR, 1.68) and the recent ecological climb agree in direction, but the earlier decline warns against a simple burden narrative; the composition of PAD deaths reflects competing influences, including changes in diabetes care [18], in the age structure of decedents, and in the coding of the underlying cause, and the data at hand cannot decompose these. Early cohort work linked diabetes to amputation and death [19], and recent national data confirm excess cardiovascular risk and mortality after amputation in type 2 diabetes [20]. National amputation series show parallel evolution [21], revascularization practice has shifted as well [22], and emergency burden after Charcot neuro-osteoarthropathy adds further weight [23].

### The kidney bridge: a designed discordance

Chronic kidney disease produced the one discordant cell in the matrix, and under the pre-specified conflict plan the divergence resolves as a difference in estimands rather than a contradiction of evidence. The individual-level analysis answers who develops PAD: adults with reduced kidney function carried a nearly doubled odds of PAD, a finding consistent with the clinical observation that reduced eGFR clusters with extensive atherosclerosis [24]. The ecological analysis answers whose PAD deaths accumulate: the share of PAD deaths mentioning renal failure was essentially unchanged in 26 years, under both the narrow N17-N19 definition (-0.4%, p = 0.74) and the broad N00-N99 chapter (+3.0%, p = 0.014), which accounted for 88% to 92% of chapter mentions throughout; in bootstrap terms, the probability that the narrow-definition share rose over the window was 0.445, with a 95% interval for the share difference of -0.7 to +0.5 percentage points, and the broad definition crossed the rising threshold (probability, 0.983) only with a share difference of at most 0.9 percentage points; both changes are trivially small next to the tobacco bridge. The flat bridge therefore argues against a narrative in which the rising prevalence of kidney disease [25] is driving the national PAD mortality composition upward. That narrative would have required the renal-failure share to rise, and it did not.

This discordance is not a weakness of the design; it is the reason the design exists. A reader relying on the individual-level literature alone could reasonably infer that kidney disease is an ascending force in PAD mortality. A reader relying on national mortality tabulations alone could not test that inference at all. Putting both streams on one table shows that the inference fails, and shows where the boundary of each evidence stream lies.

### What this design adds

To our knowledge, this is the first study to use individual-ecological triangulation as the primary analysis for PAD, combining examination-based individual data with national multiple-cause mortality records. The existing literature that touches both NHANES and CDC WONDER consists largely of simulation and policy-forecast studies [26]; descriptive national PAD mortality trends exist [27], as do numerous risk-factor studies, but the two have not previously been pre-specified against each other with a conflict plan.

The contribution is consequently methodological as much as substantive [28]: it demonstrates a template, prior hypothesis, pre-specified bridges, pre-specified conflict options, directional comparison quantified by bootstrap direction-agreement probabilities, that can be reused for other outcomes where the public-use mortality files lack disease-specific codes and the ecological layer is the only source of disease-specific mortality granularity. That data limitation of the NHANES linkage files is, in this design, converted from an obstacle into a rationale.

### Public health implications

The actionable signal is narrow and deliberately so. Both evidence streams point at smoking: it carried the strongest individual-level association with PAD, and its footprint on national PAD deaths widened steadily over the study window. Diabetes showed the same directional agreement, with the ecological signal concentrated after 2014.

Hypertension and kidney disease each mark elevated individual risk, but neither showed a rising share of PAD deaths, and hypercholesterolemia was not associated with PAD in any model. Resource allocation for PAD prevention at the population level, as distinct from risk management at the bedside, is therefore best anchored on tobacco control and diabetes management, with the individual-level associations of the other factors retained for clinical case-finding rather than population attribution. Smoking cessation carries direct outcome evidence in PAD [29]. For diabetes, incretin-based metabolic therapy is generating new outcome data in PAD populations [30]. Lipid management remains guideline-supported even where total cholesterol alone fails to discriminate [31], and adherence to proven antithrombotic therapy is an emerging system-level priority [32]. Guideline-directed care spans supervised exercise [33] and continues to be tested with novel metabolic agents [34], while performance-measure frameworks now track the delivery of these interventions [35].

### Limitations

Several limitations bound these conclusions. First, attributable fractions are formula-dependent: Levin and Miettinen estimates differed by up to 12 percentage points and did not preserve their ordering, so we report only the qualitative conclusion that smoking contributes one of the largest attributable shares, with both sets of estimates in the supplementary material. Second, the tobacco bridge is affected by the 2003 coding-rule change, and its pre-2005 segment understates mentions. Third, the ecological layer is compositional: a rising or falling share of mentions does not identify causal contributions, and the absence of individual covariates in death records leaves cross-level confounding formally possible, which is why the design claims direction only. Fourth, the individual-level exposure was measured in 1999-2004 while the ecological window extends to 2024, so the individual estimates describe a cohort whose subsequent exposures the data cannot track. Fifth, the individual mortality layer rests on a finite number of events, 235 cases of prevalent PAD and 388 cardiovascular deaths, so the individual-level hazard estimates carry correspondingly wide confidence intervals; the public-use mortality files additionally provide leading-cause groups rather than ICD-specific causes, which is precisely why the individual layer cannot answer disease-specific mortality questions and the ecological layer was brought in. Sixth, all data are from the United States, and the coding behavior of other jurisdictions may differ.

## Conclusion

Taken together, the two evidence streams behave as a triangulation design should. Where they agreed, on smoking and diabetes, each stream covers a weakness of the other: the individual estimates carry the causal interpretation and the covariate control, while the national series shows that the same factors are visibly reshaping PAD mortality at the population scale across a quarter century. Where they diverged, on kidney disease, the divergence is itself informative rather than contradictory: a cross-sectional association answers who develops PAD, whereas a compositional trend answers whose PAD deaths accumulate, and the second question was answered with a firm no. Smoking and diabetes are the two factors for which individual-level risk and population-level mortality composition agree in direction over a quarter century; kidney disease marks individual risk without moving the national mortality composition. Individual-ecological triangulation, applied here for the first time as the primary analysis for PAD, converts two incomplete evidence streams into one interpretable pattern, with the discordant cell doing as much explanatory work as the concordant ones. The practical implication is narrow but well supported: smoking cessation and diabetes control, already first-line targets in PAD care [36], are the two factors for which both the individual evidence and the population mortality record point in the same direction; kidney disease, by contrast, marks elevated PAD risk at the bedside without evidence that it is driving the national PAD death burden upward.

## Clinical Perspective

Peripheral artery disease (PAD) affects 5.05% of US adults aged 40 years and older and carried a near-threefold risk of cardiovascular death in this nationally representative cohort. Individual-level cohorts and national mortality statistics are usually cited side by side, but they have not previously been formally compared within one design. This study triangulates the two evidence streams for PAD with three pre-specified comorbidity bridges and a pre-specified conflict plan, and adds a bootstrap direction-agreement probability to each bridge. The clinical perspective is as follows:

- Current smoking and diabetes are the two risk factors supported by both evidence streams: each was independently associated with prevalent PAD at the individual level, and the share of national PAD deaths mentioning each factor rose over 26 years, from 2.20% to 18.50% for tobacco and from 7.35% to 10.92% for diabetes after its 2014 trough.
- Chronic kidney disease and hypertension mark elevated individual PAD risk but did not move the national mortality composition; hypercholesterolemia measured as total cholesterol was not associated with PAD in any model.
- Population-level PAD prevention is best anchored on tobacco control and diabetes management, while the remaining associations are better reserved for clinical case-finding at the bedside.

## Declarations

### Data availability

All data used in this study are publicly available and free of charge: NHANES 1999-2004 examination, laboratory, and questionnaire files and the NCHS linked mortality files are distributed by the National Center for Health Statistics (https://www.cdc.gov/nchs/nhanes/), and the Multiple Cause of Death files for 1999-2024 are distributed through CDC WONDER (https://wonder.cdc.gov/). The analysis code that reproduces all reported estimates is available from the corresponding author on reasonable request.

### Ethics statement

This study is a secondary analysis of publicly available, de-identified data: the NHANES 1999-2004 examination and linked mortality files released by the National Center for Health Statistics, and the aggregated Multiple Cause of Death records released through the CDC WONDER system. Under 45 CFR 46.104, exemption 4, research using such pre-existing, publicly released, non-identifiable data does not constitute human subjects research and does not require institutional review board approval or individual informed consent. Use of the NCHS linked mortality files followed the published data use restrictions, and no cell based on fewer than 10 deaths is reported anywhere in this article.

### Author contributions

Ying Zhang: methodology, validation, interpretation of the results, writing of the original draft. Jianjun Shi: supervision, project administration, writing, review and editing. Rong Wu: data curation, validation, writing, review and editing. Fei Zhao: conceptualization, methodology, software, formal analysis, data curation, visualization, writing, review and editing, supervision, and final approval of the version to be published. All authors read and approved the final manuscript.

### Declaration of generative AI use in scientific writing

During the preparation of this work, the authors used a large language model writing assistant (WorkBuddy) for language polishing, reference formatting, and text layout. No artificial intelligence tool was used to generate, simulate, or modify research data; all analyses were defined and executed by the authors and independently checked. The authors reviewed and edited all AI-assisted output and take full responsibility for the integrity of the data, the accuracy of the analysis, and the content of the published article.

